# The Implementation and Evaluation of the James Hakim Leadership Development Program in Africa: process, lessons Learned, and Feedback from the Participants

**DOI:** 10.1101/2023.07.07.23292340

**Authors:** Aloysius Gonzaga Mubuuke, Alyssa Bercasio, Georgina Yeboah, Elsie Kiguli-Malwadde, Abigail Kazembe, Maeve Forster, Deborah von Zinkernagel, Ellie Anderson, Clara E. Sam-Woode, Oathokwa Nkomazana, Patricia Katowa Mukwato, Michael J A Reid, Marietjie de Villiers

**Author notes:** Contributed equally Affiliations (Michael J A Reid, Marietjie de Villiers). Corresponding Author: Aloysius Gonzaga Mubuuke, School of Medicine, College of Health Sciences, Makerere University, Uganda.

## Abstract

**Background:** Effective leadership is crucial for improving the quality of health professionals trained in Sub-Saharan Africa (SSA). However, many health professions training institutions lack formal faculty mentorship programs for leaders, leaving faculty to learn on the job without formal support. To address this gap, the African Forum for Research and Education in Health (AFREhealth) developed an innovative leadership capacity-strengthening program, named after the late educator and research, James Hakim. Objective: This article describes the design and implementation of the AFREhealth leadership training program and how it could bridge the leadership skills gap in health professions education in SSA. The objective of the article is to describe the program’s implementation process, share the experiences of participants, and discuss lessons learned.

**Methods:** The AFREhealth leadership training program was developed through consultative meetings, after a landscape review of existing leadership training programs. The program was designed to be delivered virtually over a 12-week period, and the curriculum included modules on leadership styles and personality, mentorship, change management, conflict management, budgeting, resource mobilization, building partnerships, inter-professional education & collaborative practice, and working on inter-professional teams. Training activities included weekly workshops, small group discussions, readings, reflective sessions with senior health leaders/experts, focused mentorship sessions, and a guided project design capstone. Surveys were conducted to obtain feedback from participants and assess the program’s impact on their ongoing leadership roles in their institutions.

**Results:** The leadership training program was implemented twice in a 20-month period, with 68 trainees completing the program. Participants reported increased knowledge, skills, and confidence in attaining key leadership competencies. The virtual delivery of the training allowed for a wide pool of applicants to participate, and the curriculum was designed to be adaptable for other institutions.

**Conclusion:** The AFREhealth leadership development program demonstrated the need for mentoring health professions education leaders in Africa and the effectiveness of virtual training methods. The innovative curriculum and delivery model provide a valuable resource for other institutions seeking to build leadership capacity in health professions education.

## Background

In recent years, there has been substantive progress in advancing Health Professions Education (HPE) across Sub-Saharan Africa (SSA), given recognition of the importance of expanding the health workforce to respond to diverse clinical and public health challenges. There has been increased focus on the critical role of HPE research,^1, 2^, and effective interprofessional collaboration is essential to that advancement.^2–4^ In addition, transformative and effective leadership in HPE is also essential to ensuring the quality of training for health professionals in SSA. Effective HPE leadership is necessary to drive innovation in curriculum design, scale training programs, and respond to new and emerging public health and clinical priorities. Faced with numerous ongoing challenges, including long-standing communicable disease threats,^5, 6^ rising prevalence of non-communicable diseases,^7^ and ongoing workforce constraints,^8^ HPE leaders at all levels of the health system need to steer their institutions to adopt and adapt to evolving science and exogenous stressors. As illustrated during the COVID-19 pandemic, effective HPE leadership in Sub-Saharan Africa is especially important to how the health workforce responds to emerging public health threats.^6^

Existing health professions leadership training programs in SSA^9–11^ tend to target upper-level faculty who are already in leadership positions and are more often targeted to physicians than nurses or other health cadres. These training programs tend to be skewed toward equipping health researchers and public health practitioners, rather than health educators.^10, 12–14^ However, leadership is not only confined to individuals in formal leadership positions nor to those pursuing health research; health professions educators at all levels meet challenges that require appropriate leadership skills. Junior and mid-career faculty in African health professions training institutions, especially nurses and midwives, often lack training in key leadership skills. To ensure the sustainability of high-quality health professions education programs, it is necessary to build the next generation of leaders across various health professions education training institutions. However, leadership training programs for academic health professions training schools that train early and mid-career academic professionals with an inter-professional focus are scarce.^15^ Through leadership training of early and mid-career faculty using an inter-professional educational approach, there will be sufficient skilled professionals to create change and sustain innovation across health systems.

The lack of HPE-focused leadership training opportunities for faculty across African institutions prompted the launch of the James Hakim Leadership Development Program (JHLDP) in April 2021. This paper aims to present the implementation and evaluation of the program, which targeted early and mid-level faculty in African HPE institutions. By sharing our insights, we hope to provide valuable lessons for other institutions planning similar initiatives.

## Methods

### Setting

**African Forum for Research and Education in Health (**AFREhealth) is an interdisciplinary health organization that improves healthcare quality and health professions training in Africa through education, research, and capacity building. AFREhealth provided the operational platform for conducting the leadership training.^16, 17^

### Needs Assessment

Given the paucity of Africa-based literature on leadership training programs targeting those engaged in HPE in SSA, we first surveyed health professionals from a subset of African Health Professions (HP) training institutions affiliated with the AFREhealth network. None of those surveyed reported any formal leadership training opportunities targeting HPE at their institutions. We also conducted key informant interviews with diverse individuals engaged in health professions education from across SSA, as well as members of the AFREhealth network. These interviews highlighted key themes including lack of training in change management, program design, resource mobilization, interprofessional education, mentorship, leadership styles, project management, budgeting and conflict resolution.

### Curriculum Development

Informed by the survey and key informant interviews, stakeholders from across the AFREhealth network were invited to participate in the design of the JHDLP. Consultative meetings occurred between March and June 2021, during which time the core curriculum and recruitment strategies were developed. Seven core modules were created, informed by an evidence-based stepwise process (see Table 1).^18^ For each module, goals, objectives, and expected outcomes were determined. Then educational strategies and evaluation tools were applied to match the goals and outcomes. All content was developed to optimize interprofessional engagement since it was anticipated that emerging leaders from across diverse health professions cadres would participate. All activities were designed for online learning rather than in-person instruction due to the COVID-19 social distancing practices.

**Table 1.** Modules of the Curriculum

|  |
| --- |
| **Module 1:** Understanding Leadership Styles |
| **Module 2:** Change Management and Sustaining Change |
| **Module 3:** Project Design |
| **Module 4:** Mentorship |
| **Module 5:** Budgeting, Resource Mobilization & Partnership Development |
| **Module 6:** Conflict Management |
| **Module 7:** Inter-professional Education & Working in Inter-professional teams |

### Implementation

Both didactic lectures and small group discussion activities leveraged the Zoom platform, each occurring bi-weekly over a three-month period. In addition to these group-based sessions, the fellowship required participants to meet with a mentor with whom they were matched based on clinical interest or technical expertise. The course also included four interactive discussions with senior health leaders from across Africa. These informal sessions offered participants the opportunity to glean insights on leadership and career development from health leaders at the end of their careers.

All participants were required to design a programmatic, educational intervention, or evaluation project that they could implement after the end of the course. Monthly project progress on the write-up of these proposals were required, and at the end of the training, fellows were required to submit a complete proposal for a project they might undertake post-fellowship.

Recruitment of eligible fellowship candidates was undertaken two months prior to the launch of each course. Eligible candidates were required to be early-or mid-career faculty engaged in HPE, working at AFREhealth-affiliated academic institutions, with letters of support from academic leadership at those institutions, and evidence of professional commitment to excellence in HPE. A formal faculty position was not a prerequisite to participation, although all were expected to be clinical instructors engaged with pre-service and in-service education of health professionals. An orientation webinar was conducted for the selected faculty, mentors, and trainers to familiarize themselves with the program’s objectives and expected outcomes prior to the launch of the course.

### Evaluation

Prior to participation and on completion of the training, an online survey was conducted to gather the views and experiences of the participants (faculty, trainers, and mentors) and to assess the confidence of fellows in leadership domains relevant to the curriculum. Fellows were invited to complete an additional online survey >12 months after participation in the program to assess the impact of the course on subsequent professional practice, including evaluating the extent to which skills and competencies addressed in the program were subsequently employed in their professional work. The survey was anonymous, and investigators had no way of identifying respondents during or after data collection. The collected data were coded and analyzed. Descriptive analysis was performed for quantitative data, and thematic analysis was used for qualitative data to identify common patterns across responses.

### Ethics

The study received an ethics waiver from the School of Medicine Research Ethics Committee at Makerere University as was deemed zero-risk. Verbal consent to participate in the feedback survey was requested of all participants at the start of each cohort. All evaluations of fellows, facilitators, and mentors were anonymous.

## Results

Between January and May 2021, this comprehensive HPE leadership development program was developed to equip emerging leaders in HPE across SSA. Each module was delivered in an interprofessional format to address the critical competencies of HPE leaders, regardless of health professions cadre. **Table 1** summarizes the core modules addressed in the program.

Across the two cohorts of learners, the first recruited prior to June 2021 and the second recruited prior to June 2022, 124 applicants applied from 16 countries and 25 health professions training institutions. In 2021, 40 fellows were enrolled in the JHDLP. The following year, 28 fellows were enrolled. As outlined in Table 2, 32% (n=23) of these fellows were nurses or midwives, and 43% (n=29) were physicians. **Table 2** highlights the cadres represented in the leadership training program as well as their demographic characteristics. Across both cohorts, 100% (n=68) of participants completed both the pre-and post-course assessments, and 49% (n=33) of all candidates completed the final post-participation survey, at a point >12 months after participation in the initial course.

**Table 2.**
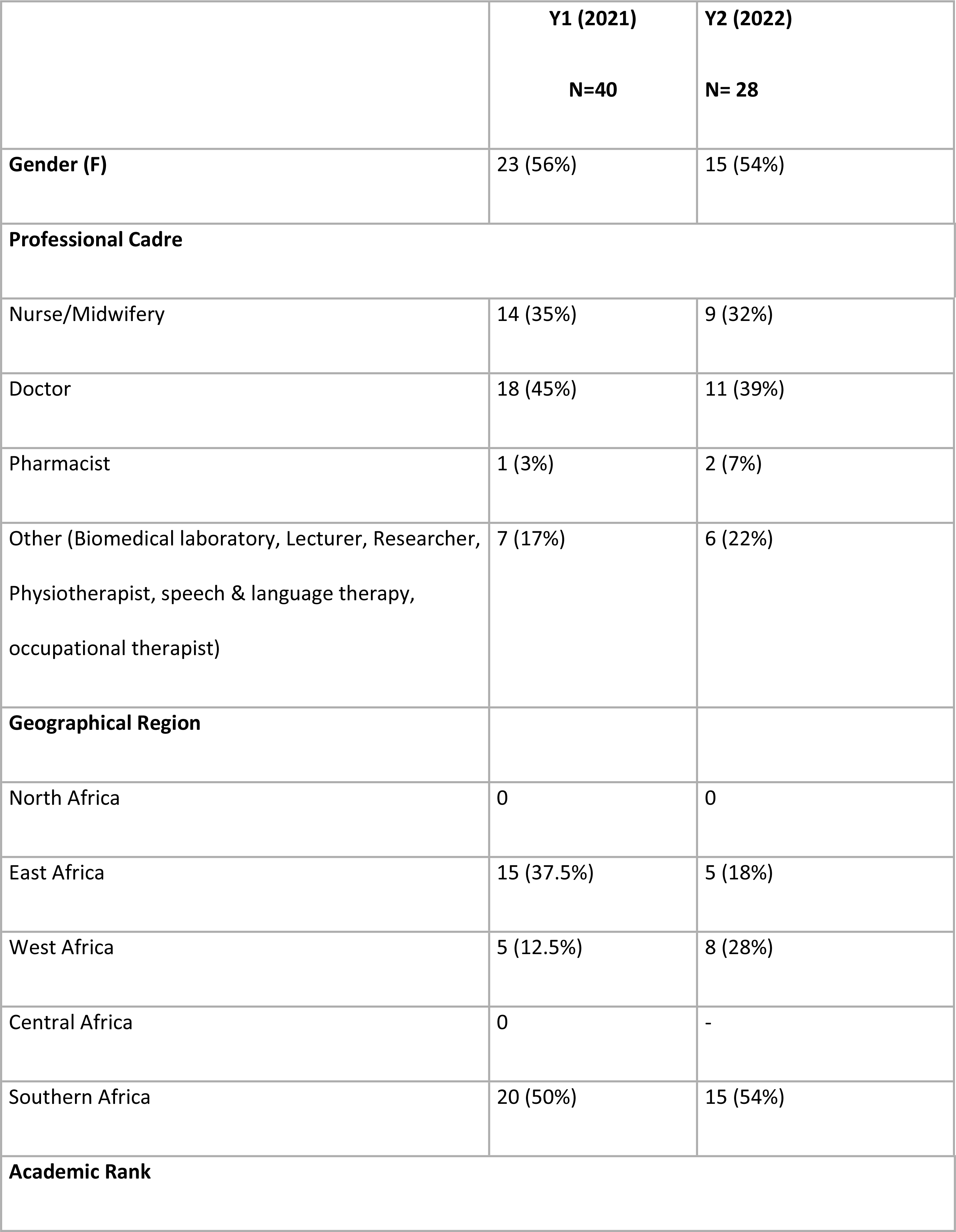
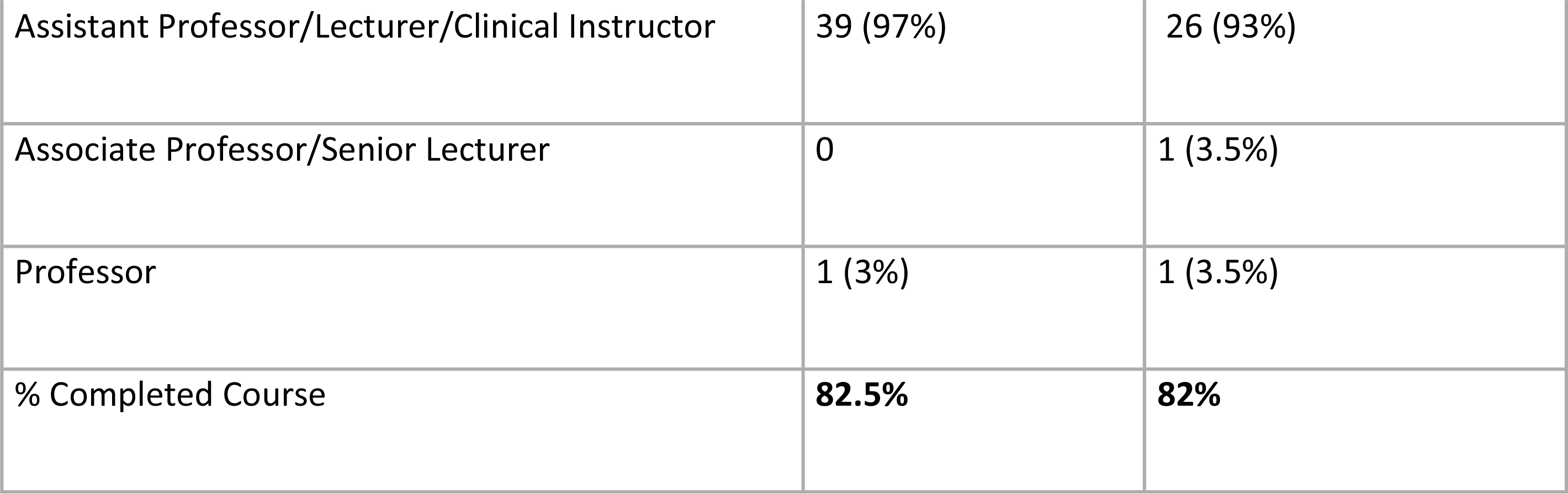
Participant Demographics.

|  | **Y1 (2021)**<br>**N=40** | **Y2 (2022)**<br>**N= 28** |
| --- | --- | --- |
| **Gender (F)** | 23 (56%) | 15 (54%) |
| **Professional Cadre** |  |  |
| Nurse/Midwifery | 14 (35%) | 9 (32%) |
| Doctor | 18 (45%) | 11 (39%) |
| Pharmacist | 1 (3%) | 2 (7%) |
| Other (Biomedical laboratory, Lecturer, Researcher, Physiotherapist, speech & language therapy, occupational therapist) | 7 (17%) | 6 (22%) |
| **Geographical Region** |  |  |
| North Africa | 0 | 0 |
| East Africa | 15 (37.5%) | 5 (18%) |
| West Africa | 5 (12.5%) | 8 (28%) |
| Central Africa | 0 | - |
| Southern Africa | 20 (50%) | 15 (54%) |
| **Academic Rank** |  |  |
| Assistant Professor/Lecturer/Clinical Instructor | 39 (97%) | 26 (93%) |
| Associate Professor/Senior Lecturer | 0 | 1 (3.5%) |
| Professor | 1 (3%) | 1 (3.5%) |
| % Completed Course | **82.5%** | **82%** |

From those who completed the final post-participation survey, and as illustrated in Fig.1 the majority reported that the training on interprofessional education practice (24, 73%), leadership style (21, 64%), and effective mentorship (22, 67%) were extremely useful to their professional practice. Training on budget management and resource mobilization and partnership development was deemed to be extremely useful by fewer participants (55% (n=18) and 58% (n=19), respectively).

**Fig. 1.**
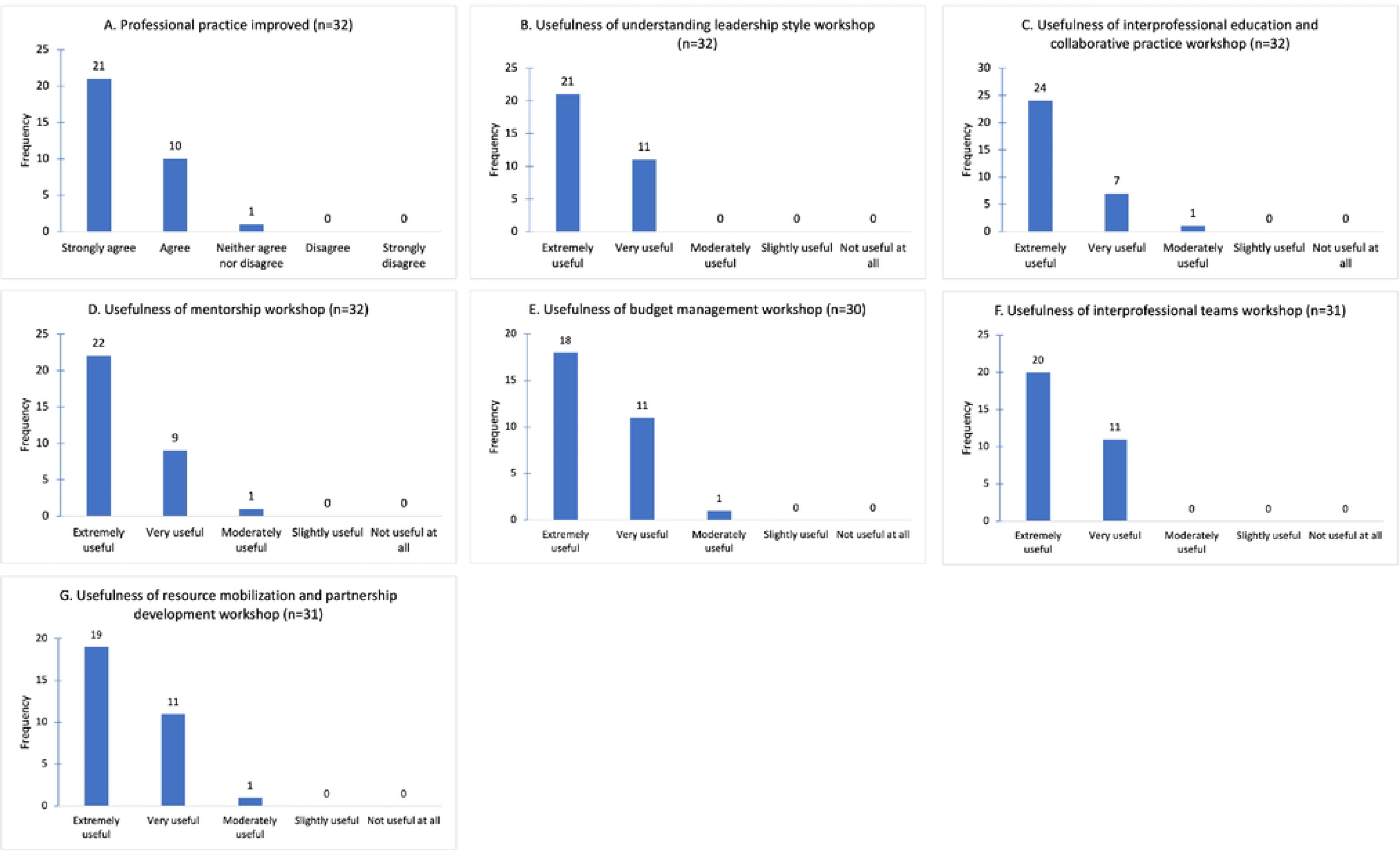
Feedback from participants in the Leadership Training Program, (feedback received >12 months post participation)

The majority of those who completed the alumni survey >12 months after participation in the course (97%, n=32) stated that their leadership responsibilities had expanded since participation in the course: respondents detailed that they had since undertaken new educational (19, 57.6%), research (28, 84.8%), or administrative (19, 57.6%) leadership responsibilities. All respondents also reported increased motivation to engage in more multidisciplinary approaches to health professions education leadership in the future within their home institutions.

## Discussion

Effective leadership is crucial for improving the quality of health professionals trained in sub-Saharan Africa, and the lack of formal faculty mentorship programs for leaders has left many early career professionals to learn these skills without formal support.^9, 10^ This study demonstrated how a leadership program targeting emerging HPE leaders in SSA was successfully implemented and provided participants with valuable knowledge, skills, and confidence in attaining key leadership competencies. To our knowledge, the JHDLP is also the first program for emerging HPE leaders in SSA. Our results also highlight JHDLP’s sustained impact; most fellows reported the training continued to have positive impacts on their career even >12 months after participation, equipping them with skills not only to respond to HPE challenges in their settings but also to address unique threats posed by the COVID-19 pandemic. Outlined below are key lessons learned from the program.

First, the JHDLP leveraged virtual learning modalities, enabling participants to access the training remotely. This was particularly useful because the COVID-19 pandemic made it challenging to gather in person for training activities. The virtual delivery of the training also enabled a diverse range of fellows to participate, including nurses and midwives engaged in frontline training programs in remote or rural settings. These results affirm the critical role that online training interventions can play in advancing professional development opportunities, especially for health professions leaders, including those historically underserved by training initiatives such as nurses and midwives, delivered in a format that is also likely to be less expensive and disruptive than in-person training programs.^7, 19^

Second, the JHDLP highlights the importance of interprofessional training as critical to both leadership development and effective interprofessional collaboration. By integrating diverse perspectives and knowledge, interprofessional leadership training initiatives like this one can equip emerging leaders with the competencies needed to tackle complex challenges that many academic HPE leaders face managing competing academic, administrative and educational priorities. Moreover, the ability to collaborate with professionals from different backgrounds and disciplines is likely to enhance their capacity to design comprehensive and well-rounded educational curricula. This, in turn, may lead to improved learning experiences for students and a higher quality of education overall. In addition, the benefits of interprofessional leadership training extend beyond educational programs. Leaders who can effectively collaborate with professionals from different fields are also better equipped to oversee healthcare delivery. They are more likely to understand the importance of interdisciplinary teamwork and foster an environment that promotes effective communication, collaboration, and coordination among healthcare providers.

Third, the JHDLP’s implementation during a pandemic highlights the utility of this kind of intervention to prepare leaders in the midst of, and in preparation for, public health and clinical crises. The virtual format enabled participants to acquire crucial training even though they were geographically disbursed and unable to meet in person. Moreover, this program included a focus on equipping participants with relevant skills to ensure operational excellence, technical expertise, and diplomatic competency necessary for managing clinical crises and challenges such as the pandemic.

There are some limitations to this study. Our analysis was based solely on the feedback received from those who participated and completed the feedback survey. Only 49% of participants completed the alumni survey and we are cautious to make inferences about how those who did not complete this follow-up survey perceived its utility; we acknowledge that important information could have been missed from those who did not complete the feedback survey. Moreover, this is only an evaluation to obtain a snapshot of what the participants experienced during the program. It does not necessarily measure the real impact in the various institutions where the faculty come from. The impact of the fellowship program on clinical and educational outcomes is the subject of a future report which will include an assessment of the participant’s knowledge, and confidence and will be based on narratives, reflections, and focus group discussions that are ongoing.

### Conclusion

The James Hakim Leadership Development Program demonstrated the effectiveness of a virtual training program and the need for mentoring HPE leaders in Africa. The innovative curriculum and delivery model provides a valuable resource for other institutions seeking to build leadership capacity in health professions education. Future research should expand the program to larger samples and evaluate the impact of the training on trainees’ professional development and the quality of healthcare provision in the region.

## Data Availability

All data is included in the manuscript.

## Acknowledgment & Funding

This study was funded by AFREhealth with support from UCSF and HRSA

